# Characteristics and care trajectories of older patients in temporary stays in Denmark

**DOI:** 10.1101/2024.10.10.24315219

**Authors:** Hanin Harbi, Carina Lundby, Peter Bjødstrup Jensen, Søren Post Larsen, Linda Grouleff Rørbæk, Lene Vestergaard Ravn-Nielsen, Jesper Ryg, Mette Reilev, Kasper Edwards, Anton Pottegård

**Author notes:** Correspondence: Anton Pottegård, Clinical Pharmacology, Pharmacy and Environmental Medicine, Department of Public Health, University of Southern Denmark, Campusvej 55, 5230 Odense M, Denmark.

## Abstract

**Background:** Temporary stays for patients requiring short-term care outside the home, often following hospital discharge, has gained increasing importance. This study aimed to describe the characteristics and care trajectories of older patients in Danish temporary stays to improve care delivery and patient safety.

**Methods:** We conducted a descriptive study on a cohort of patients in temporary stays across 14 Danish municipalities from 2016 to 2023, using data from national health registries.

**Results:** We identified 11,424 patients with a median age of 81 years (interquartile range [IQR] 73-87 years); 54% were women. Patients exhibited a high level of comorbidity, with a median Charlson Comorbidity Index of 1 (IQR 0-2), and a median of 3 hospital admissions (IQR 2-6) in the year preceding their move into temporary care. The majority (70%) transitioned to temporary stays following hospital discharge, while 30% were admitted directly from their homes. The median duration of temporary stays was 24 days (IQR 11-49 days), with 9.1% staying ≥ 90 days. Additionally, 8.2% of patients were hospitalised directly from the temporary stay facility, with a median time to hospital admission of 13 days (IQR 5-28 days). Median survival after admission to a temporary stay was 23 months (IQR 3.6-57 months). Predictors of mortality included male sex, older age, higher comorbidity burden, and increased number of hospital admissions prior to temporary stay.

**Conclusion:** Patients in temporary stays are generally older individuals with multimorbidity and limited life expectancy. Most patients are admitted following hospital discharge, and their stays are often prolonged.

## Introduction

As the life expectancy increases, and population ages, more people are living with chronic diseases and disabilities. This demographic shift places significant strain on healthcare systems, which again has in most settings led to earlier hospital discharges of patients with more complex and fluctuating care needs. To manage these accelerated discharges and reduce hospital (re)admissions, many countries have reorganized their healthcare systems to strengthen patient transitions between different levels of care [1,2]. Globally, various terms describe these transitional services, such as “intermediate care” in Europe, “subacute care”, “postacute care”, or “skilled nursing care” in the United States, “transition care” in Australia, and “transitional care” in Canada [3–5]. Intermediate care services are designed to provide short-term care to individuals who are discharged from the hospital but are not yet ready to return home, or who are at risk of hospital admission. These services can either be home-based or bed-based, with the latter involving temporary stays at designated care facilities [2,6–8].

In Denmark, these bed-based intermediate care services are referred to as “temporary stay facilities”. Temporary stays have become increasingly important in the Danish healthcare system, especially for older patients with frailty, multimorbidity, and complex medication regimens. However, the complexity of the patients’ conditions poses significant challenges for healthcare staff, who may not always have the necessary resources or expertise to meet their specialized needs [9–14].

To improve the care and safety of patients in temporary stays, it is essential to gain deeper understanding of their characteristics and care trajectories. The aim of this study was to provide a detailed description of the characteristics, health profiles, and outcomes of patients in temporary stays in Denmark.

## Methods

We established a cohort comprising 14,978 temporary stays from 11,424 patients across 14 Danish municipalities between 2016 and 2023. The cohort was supplemented with individual-level data from Danish national administrative and health registries to describe the characteristics and trajectories of patients in temporary stays.

### Data sources

The municipalities provided data on temporary stays from January 1, 2016, to December 31, 2023, including the patient’s move-in date and move-out date, along with their Central Person Register (CPR) number, a unique personal identifier assigned by the Civil Registration System to all Danish residents since 1968 [15]. These data were linked to nationwide health registries using the CPR number. Information on hospital admission and diagnoses was obtained from the Danish National Patient Registry, which contains data on all nonpsychiatric hospital admissions since 1977 and both psychiatric and nonpsychiatric outpatient contacts since 1995. Diagnoses have been coded according to the 10^th^ revision of the International Classification of Diseases (ICD-10), from 1994 onwards [16]. We retrieved demographic data (age, sex, death, and migration) from the Civil Registration System [15]. For comorbidity assessment, prescription drug use was obtained from the Danish National Prescription Registry, which contains records of all prescription drugs dispensed by Danish community pharmacies since 1995 [17,18]. We obtained information on care home admissions from a nationwide cohort of care home admissions maintained by the Danish Health Data Authority, covering admissions from 2015 onwards.

### Study cohort

Temporary stays were included if both the move-in and move-out dates occurred within the study period. We excluded temporary stays with missing or invalid CPR numbers, move-in dates, or move-out dates and those where the move-out date preceded the move-in date. Patients were required to have resided in Denmark for at least two years prior to their first temporary stay. For individuals with multiple temporary stays, consecutive stays with no gap between move-in and move-out dates were combined into a single continuous stay. Only the first temporary stay for each patient was included in the analyses.

### Setting

In Denmark, temporary stays are provided by municipalities for individuals requiring short-term care and support that cannot be managed at home. These stays can be used for care, treatment, or rehabilitation following an illness or hospitalisation, as well as for providing respite care for family caregivers. Access to temporary stays is managed by the municipalities, and the services and organisation can vary across them. Care is delivered by multidisciplinary teams, including nurses, care assistants, physiotherapists, and occupational therapists. Temporary stay facilities are not required to have physicians on staff. Instead, medical responsibility remains with the patient’s general practitioner (GP) or the hospital [11].

### Analyses

We conducted a series of analyses to describe the study cohort and patient trajectories. First, we described patient characteristics at the time of move-in overall and stratified by sex and age group (< 75, 75-84, and ≥ 85 years). Baseline characteristics included sex, age, Charlson Comorbidity Index [19], selected comorbidities (Appendix A), and number of hospital admissions in the year prior to move-in. The Charlson Comorbidity Index and comorbidities were derived from ICD-10 hospital discharge diagnoses and prescription data from the Danish National Patient Registry and Danish National Prescription Registry, using data from 10 years before baseline.

Second, we examined patient locations prior to move-in by categorizing admissions into temporary stays as following hospital discharge (defined as discharge on or the day before move-in date), from home, or from a care home, both overall and by municipality. For hospital admissions leading to temporary stays, the primary diagnosis was identified. Additionally, we calculated the median length of stay overall, by municipality, and by patient location prior to move-in. We assessed destinations after move-in by examining the proportion of patients hospitalised directly from the facility (on the move-out date or the day after), within 30 days of move-in or move-out, and the proportion of patients transferred to a care home within 30 days of move-out. These analyses included only surviving patients. A supplementary analysis extended the window to 90 days. For hospital admissions from temporary stay facilities, we assessed the median time to hospital admission, the timing of admission by hour and the day of the week, and the primary reason for admission, overall and by weekdays and weekends.

Third, to describe the mortality of patients in temporary stays, we estimated median survival after move-in and 30-day, 90-day, and 1-year survival rates, overall and by sex and age group, using the Kaplan-Meier method. We calculated odds ratios (ORs) for 30- and 90-day mortality predictors using logistic regression, with sex, age, Charlson Comorbidity Index, selected comorbidities (Appendix A), and hospital admissions in the year prior to move-in included as potential predictors.

All statistical analyses were conducted using R version 4.3.3.

### Data and ethics

The study was registered at the University of Southern Denmark’s inventory (record no. 11.436). Ethical approval is not required for registry-based studies in Denmark.

## Results

We identified 14,978 temporary stays from 11,424 patients during the study period, including only the first temporary stay for each patient in our analyses (Table 1). More than half of the patients (54%) were women. The median age at move-in was 81 years (interquartile range [IQR], 73-87 years), with women being slightly older than men (median age 83 versus 79 years, p < 0.001). The median Charlson Comorbidity Index was 1 (IQR 0-2) and patients had a median of 3 hospital admissions (IQR 2-6) in the year prior to move-in.

**Table 1.** Baseline characteristics of patients moving into temporary stay facilities in 14 Danish municipalities from 2016 to 2023, overall and stratified by sex and age groups.

|  | Total<br>(n = 11,424) | Women<br>(n = 6,141) | Men<br>(n = 5,283) | < 75 years<br>(n = 3,394) | 75-84 years<br>(n = 4,017) | ≥ 85 years<br>(n = 4,013) |
| --- | --- | --- | --- | --- | --- | --- |
| Sex |  |  |  |  |  |  |
| Female | 6,141 (54%) | 6,141 (100%) | 0 (0.00%) | 1,470 (43%) | 2,120 (53%) | 2,551 (64%) |
| Male | 5,283 (46%) | 0 (0.00%) | 5,283 (100%) | 1,924 (57%) | 1,897 (47%) | 1,462 (36%) |
| Age |  |  |  |  |  |  |
| Median (IQR) | 81 (73-87) | 83 (75-89) | 79 (71-86) | 68 (62-72) | 80 (78-83) | 90 (87-93) |
| < 75 years | 3,394 (30%) | 1,470 (24%) | 1,924 (36%) | 3,394 (100%) | 0 (0.00%) | 0 (0.00%) |
| 75-84 years | 4,017 (35%) | 2,120 (35%) | 1,897 (36%) | 0 (0.00%) | 4,017 (100%) | 0 (0.00%) |
| ≥ 85 years | 4,013 (35%) | 2,551 (42%) | 1,462 (28%) | 0 (0.00%) | 0 (0.00%) | 4,013 (100%) |
| Charlson Comorbidity Index (CCI) |  |  |  |  |  |  |
| Median (IQR) | 1 (0-2) | 1 (0-2) | 2 (0-3) | 1 (0-3) | 2 (0-3) | 1 (0-2) |
| 0-1 | 5,752 (50%) | 3,338 (54%) | 2,414 (46%) | 1,712 (50%) | 1,948 (48%) | 2,092 (52%) |
| 2-3 | 3,921 (34%) | 2,063 (34%) | 1,858 (35%) | 1,078 (32%) | 1,441 (36%) | 1,402 (35%) |
| ≥ 4 | 1,751 (15%) | 740 (12%) | 1,011 (19%) | 604 (18%) | 628 (16%) | 519 (13%) |
| Medical history of |  |  |  |  |  |  |
| Cancer | 3,135 (27%) | 1,548 (25%) | 1,587 (30%) | 884 (26%) | 1,172 (29%) | 1,079 (27%) |
| Chronic obstructive pulmonary disease | 3,676 (32%) | 2,072 (34%) | 1,604 (30%) | 1,171 (35%) | 1,379 (34%) | 1,126 (28%) |
| Dementia | 1,428 (12%) | 746 (12%) | 682 (13%) | 254 (7.5%) | 635 (16%) | 539 (13%) |
| Parkinson disease | 477 (4.2%) | 193 (3.1%) | 284 (5.4%) | 120 (3.5%) | 248 (6.2%) | 109 (2.7%) |
| Ischemic heart disease | 6,326 (55%) | 3,215 (52%) | 3,111 (59%) | 1,515 (45%) | 2,357 (59%) | 2,454 (61%) |
| Heart failure | 4,843 (42%) | 2,601 (42%) | 2,242 (42%) | 1,209 (36%) | 1,710 (43%) | 1,924 (48%) |
| Atrial fibrillation | 2,842 (25%) | 1,382 (23%) | 1,460 (28%) | 464 (14%) | 1,086 (27%) | 1,292 (32%) |
| Stroke | 3,015 (26%) | 1,467 (24%) | 1,548 (29%) | 922 (27%) | 1,169 (29%) | 924 (23%) |
| Diabetes mellitus | 2,594 (23%) | 1,151 (19%) | 1,443 (27%) | 901 (27%) | 1,010 (25%) | 683 (17%) |
| Alcohol use disorder | 735 (6.4%) | 246 (4.0%) | 489 (9.3%) | 535 (16%) | 168 (4.2%) | 32 (0.80%) |
| Substance use disorder | 558 (4.9%) | 280 (4.6%) | 278 (5.3%) | 324 (9.5%) | 172 (4.3%) | 62 (1.5%) |
| Fall injuries | 6,499 (57%) | 3,902 (64%) | 2,597 (49%) | 1,744 (51%) | 2,190 (55%) | 2,565 (64%) |
| Hospitalizations in the year before move-in |  |  |  |  |  |  |
| Median (IQR) | 3 (2-6) | 3 (2-5) | 4 (2-7) | 4 (2-7) | 3 (2-6) | 3 (2-5) |
| 0-2 | 4,209 (37%) | 2,453 (40%) | 1,756 (33%) | 992 (29%) | 1,488 (37%) | 1,729 (43%) |
| 3-5 | 3,939 (34%) | 2,158 (35%) | 1,781 (34%) | 1,152 (34%) | 1,351 (34%) | 1,436 (36%) |
| ≥ 6 | 3,276 (29%) | 1,530 (25%) | 1,746 (33%) | 1,250 (37%) | 1,178 (29%) | 848 (21%) |

\* Charlson Comorbidity Index and medical history of comorbidities were determined using the 10<sup>th</sup> revision of the International Classification of Diseases (ICD-10) hospital discharge diagnoses and prescription records from the Danish National Patient Registry and Danish National Prescription Registry, respectively, covering the 10 years prior to move-in.

Most patients (70%) moved into a temporary stay facility following hospital discharge, while 30% came directly from home and 0.3% came from a care home. Patients entering from home had a higher prevalence of dementia (19% versus 9.4%, p < 0.001) and Parkinson disease (6.1% versus 3.4%, p < 0.001) but a lower median number of hospital admissions in the prior year (2 versus 4, p < 0.001) compared to those coming from hospital (Supplementary Table 1). The distribution of where patients came from before move-in varied across municipalities (Supplementary Table 2), with the proportion of patients coming from home ranging between 10% and 58%. Among those moving in after a hospital discharge, the three most common primary diagnoses were rehabilitation (10%), hip fracture (7.9%), and pneumonia (3.9%) (Supplementary Table 3).

The median length of a temporary stay was 24 days (IQR 11-49 days), with 9.1% of patients staying for 90 days or longer (Figure 1). Patients with stays of at least 90 days were slightly younger (median age 79 versus 81 years, p < 0.001) and had lower prevalences of cancer (21% versus 28%, p < 0.001), chronic obstructive pulmonary disease (COPD) (26% versus 33%, p < 0.001), and heart failure (34% versus 43%, p < 0.001) but higher prevalences of dementia (17% versus 12%, p < 0.001) and stroke (34% versus 26%, p < 0.001) (Supplementary Table 4). The median length of stay did not vary substantially by prior location (data not shown) or by municipality (Supplementary Figure 1), except for one outlier municipality that showed a higher median length of stay, though with similar patient characteristics (data not shown).

**Figure1.**
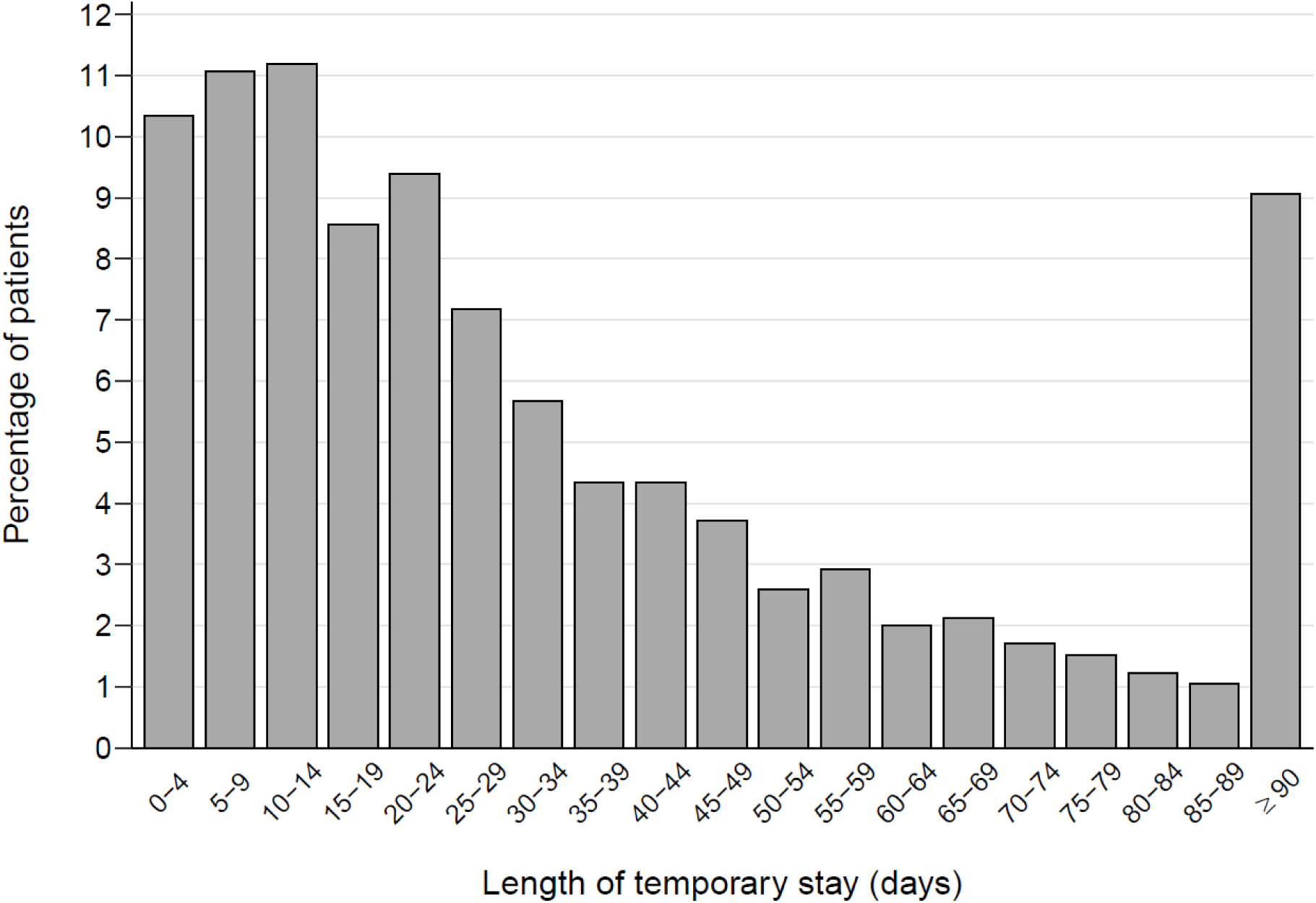
Distribution of temporary stay lengths.

Regarding patient destinations after moving out of temporary stay, 8.2% (805/9,862) were hospitalised directly from the temporary stay facility, with a median time to hospital admission of 13 days (IQR 5-28 days). Hospitalisation rates increased gradually throughout the morning, peaking at 10 a.m., and declining later in the day, with fewer admissions during evenings and nights (Supplementary Figure 2). Hospital admissions were most frequent on weekdays, particularly Mondays and Tuesdays, and least frequent on Sundays (Supplementary Figure 3). The most common reasons for hospitalisation from a temporary stay facility were pneumonia (6.8%), the need for specialised palliative care (6.1%), and radiological examination (4.2%) (Supplementary Tables 5-7). Additionally, 26% (2,522/9,865) were hospitalised within 30 days of moving into the facility, 20% (1,767/9,013) were hospitalised within 30 days of moving out, and 17% (1,495/9,013) were transferred to a care home within 30 days of moving out. Extending the window to 90 days, the corresponding proportions were 36% (3,179/8,747) for hospitalisation after moving in, 29% (2,433/8,326) for hospitalisation after moving out, and 21% (1,766/8,326) for care home transfer.

The median overall survival after moving into a temporary stay facility was 23 months (IQR 3.6-57 months). Survival rates were 86% at 30 days, 77% at 90 days, and 62% at 1 year (Figure 2). Men had shorter median survival than women (20 versus 26 months), as well as lower survival rates at 30 days (85% versus 87%), 90 days (75% versus 78%) and 1-year (59% versus 64%) (Figure 2). Survival also decreased with increasing age. The median survival was 42 months for patients under 75 years, 25 months for those aged 75-84 years, and 14 months for those 85 years and older. The 30-day, 90-day, and 1-year survival rates were 91%, 83%, and 71%, respectively, for patients under 75; 86%, 77%, and 63%, respectively, for those aged 75-84 years; and 83%, 71%, and 53% for those aged 85 or older (Figure 2).

**Figure 2.**
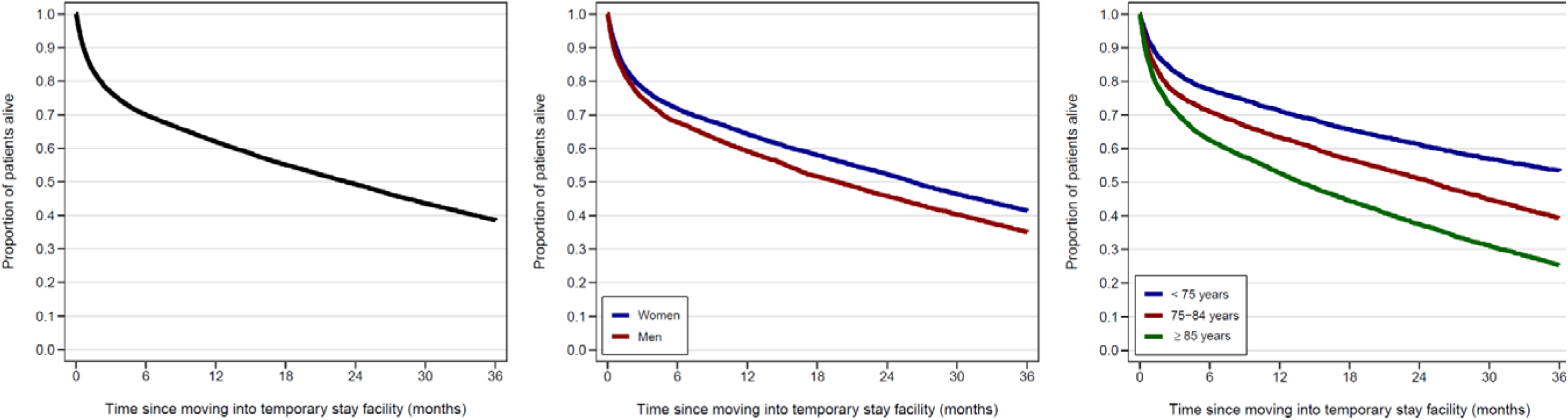
Kaplan-Meier survival curves for patients moving into temporary stay facilities in 14 Danish municipalities from 2016 to 2023, overall (left panel), stratified by sex group (middle panel), and stratified by age group (right panel)

Baseline predictors of 30-day mortality after moving into a temporary stay facility included male sex (OR 1.17, 95% confidence interval [CI] 1.05-1.32), older age (OR 1.60, 95% CI 1.37-1.87 for ages 75-84 and OR 2.29, 95% CI 1.95-2.69 for ages 85 and older compared to those under 75), higher Charlson Comorbidity Index, more hospital admissions in the year before move-in, and a history of cancer (OR 1.65, 95% CI 1.41-1.93) or heart failure (OR 1.51, 95% CI 1.34-1.71). Conversely, a history of Parkinson disease (OR 0.58, 95% CI 0.40-0.82), dementia (OR 0.72, 95% CI 0.59-0.88), and fall injuries (OR 0.77, 95% CI 0.68-0.86) was associated with decreased 30-day mortality (Table 2). A similar pattern of baseline predictors was observed for 90-day mortality (Supplementary Table 8).

**Table 2.** Baseline predictors of 30-day mortality after moving into a temporary stay facility.

|  | OR (95% CI) |
| --- | --- |
| Sex |  |
| Female | 1.00 (ref.) |
| Male | 1.17 (1.05 - 1.32) |
| Age |  |
| < 75 years | 1.00 (ref.) |
| 75-84 years | 1.60 (1.37 - 1.87) |
| ≥ 85 years | 2.29 (1.95 - 2.69) |
| Charlson Comorbidity Index (CCI) |  |
| 0-1 | 1.00 (ref.) |
| 2-3 | 1.36 (1.15 - 1.60) |
| ≥ 4 | 1.93 (1.56 - 2.40) |
| Medical history of |  |
| Cancer | 1.65 (1.41 - 1.93) |
| Chronic obstructive pulmonary disease | 1.12 (1.00 - 1.27) |
| Dementia | 0.72 (0.59 - 0.88) |
| Parkinson disease | 0.58 (0.40 - 0.82) |
| Ischemic heart disease | 0.97 (0.86 - 1.09) |
| Heart failure | 1.51 (1.34 - 1.71) |
| Atrial fibrillation | 0.97 (0.85 - 1.10) |
| Stroke | 0.82 (0.71 - 0.93) |
| Diabetes mellitus | 0.91 (0.79 - 1.04) |
| Alcohol use disorder | 0.83 (0.62 - 1.08) |
| Substance use disorder | 0.98 (0.75 - 1.28) |
| Fall injuries | 0.77 (0.68 - 0.86) |
| Hospitalizations in the year before move-in |  |
| 0-2 | 1.00 (ref.) |
| 3-5 | 1.18 (1.02 - 1.35) |
| ≥ 6 | 1.59 (1.38 - 1.83) |

## Discussion

This study describes the characteristics and care trajectories of patients in temporary stays in Denmark. Our findings indicate that these patients are generally older adults with multiple chronic conditions, most of whom enter the temporary stay facility following a hospital admission. The median length of stay was 24 days, and many patients had limited life expectancy, with a substantial proportion being hospitalised directly from or shortly after leaving the facility.

The key strength of our study is the use of a large cohort of patients across multiple municipalities in Denmark, linked to highly valid nationwide health registries [15–18], which eliminates the risk of selection bias. Additionally, access to a national cohort of all care home admissions allowed us to identify the substantial number of patients who were transferred to a care home shortly after their temporary stays.

However, this study also has limitations. We lacked data on the specific types of temporary stays (e.g., rehabilitation versus respite stay), which prevented us from investigating differences in patient characteristics and outcomes across stay types, as well as the extent of reentries and transfers (from one type to another). Given that different types of stays target distinct groups (e.g., patients in rehabilitation stays typically come from hospitals, while those in respite stays often come from home) [11], this may explain the municipal differences in the distribution of patient locations prior to move-in. Additionally, our identification of reasons for hospital admission may not have been precise. We relied on primary diagnoses from the Danish National Patient Registry, which included nondiagnostic ICD-10 R and Z codes, while secondary diagnoses were not used due to their optional nature and potential for multiple entries per admission.

This is the first study to systematically describe a large cohort of patients in temporary stays in Denmark. Previous studies from a single Danish municipality, which focused on patients entering temporary stays after hospital discharge, reported similar findings in terms of sex, age, and comorbidity burden [20,21]. However, one study observed a slightly higher 30-day mortality (17% versus 14% in our study) [21], likely because their cohort included frailer patients discharged from geriatric departments. Internationally, studies on intermediate care units in England reported a similar distribution of sex and age and a similar burden of comorbidities but noted differences in patient origins, with a lower proportion coming from hospitals (46% versus 70%) and a higher proportion from home or care homes (51% versus 30%). They also observed shorter stays (median 17 days, IQR 5-34 days versus 24 days, IQR 11-49 days in our study) and lower 1-year mortality (28% versus 38%). Similar to our findings, poorer survival was associated increasing age, higher Charlson Comorbidity Index, and cancer [22]. In Norway, patients in municipal acute wards typically enter from home and have a median length of stay of three days. However, these intermediate care units are different, as they mainly target patients coming from their home to prevent hospital admissions by managing acute conditions with short-term stays in primary care, as opposed to hospital admissions [23–25].

We observed that a considerable proportion of patients were transferred to care homes shortly after leaving the temporary stay facility. Generally, the morbidity and mortality profile of patients in temporary stays closely resembles that of Danish care home residents, with conditions like heart failure and cancer associated with poorer survival in care home residents [26]. Interestingly, Parkinson disease, dementia, and fall injuries were associated with lower mortality in temporary stays, possibly because these conditions prompt entry into temporary stays for less acute needs compared to conditions such as heart failure or cancer. This may also explain the differences in comorbidities by stay duration and premove-in location.

In conclusion, this study provides a comprehensive overview of the morbidity and mortality of patients in temporary stays, information essential for optimizing care transitions and ensuring better outcomes for patients. The findings underscore the complexity of caring for patients in temporary stays, a challenge that will likely increase as healthcare systems face rising demands and limited resources. It is crucial to consider patients’ health status when organizing temporary stays to ensure optimal care.

## Data Availability

Data are used under license from the Danish Health Data Authority

---

**Supplementary Table 1.** Baseline characteristics of patients moving into temporary stay facilities in 14 Danish municipalities from 2016 to 2023, stratified by patient location prior to move-in (hospital or home)

|  | Hospital<br>(n = 7,985) | Home<br>(n = 3,407) |
| --- | --- | --- |
| Sex |  |  |
| Female | 4,369 (55%) | 1,752 (51%) |
| Male | 3,616 (45%) | 1,655 (49%) |
| Age |  |  |
| Median (IQR) | 81 (73-87) | 82 (74-88) |
| < 75 years | 2,465 (31%) | 918 (27%) |
| 75-84 years | 2,782 (35%) | 1,227 (36%) |
| ≥ 85 years | 2,738 (34%) | 1,262 (37%) |
| Charlson Comorbidity Index (CCI)* |  |  |
| Median (IQR) | 1 (0-2) | 2 (0-3) |
| 0-1 | 4,109 (51%) | 1,629 (48%) |
| 2-3 | 2,675 (34%) | 1,230 (36%) |
| ≥ 4 | 1,201 (15%) | 548 (16%) |
| Medical history of* |  |  |
| Cancer | 2,177 (27%) | 957 (28%) |
| Chronic obstructive pulmonary disease | 2,583 (32%) | 1,086 (32%) |
| Dementia | 752 (9.4%) | 660 (19%) |
| Parkinson disease | 269 (3.4%) | 207 (6.1%) |
| Ischemic heart disease | 4,387 (55%) | 1,918 (56%) |
| Heart failure | 3,371 (42%) | 1,462 (43%) |
| Atrial fibrillation | 2,048 (26%) | 790 (23%) |
| Stroke | 2,193 (27%) | 814 (24%) |
| Diabetes mellitus | 1,814 (23%) | 773 (23%) |
| Alcohol use disorder | 542 (6.8%) | 192 (5.6%) |
| Substance use disorder | 401 (5.0%) | 156 (4.6%) |
| Fall injuries | 4,658 (58%) | 1,822 (53%) |
| Hospitalizations in the year before move-in |  |  |
| Median (IQR) | 4 (2-6) | 2 (0-5) |
| 0-2 | 2,454 (31%) | 1,730 (51%) |
| 3-5 | 3,019 (38%) | 917 (27%) |
| ≥ 6 | 2,512 (31%) | 760 (22%) |

\* Charlson Comorbidity Index and medical history of comorbidities were determined using the 10<sup>th</sup> revision of the International Classification of Diseases (ICD-10) hospital discharge diagnoses and filled prescriptions from the Danish National Patient Registry and Danish National Prescription Registry, respectively, covering the 10 years prior to move-in.

**Supplementary Table 2.**
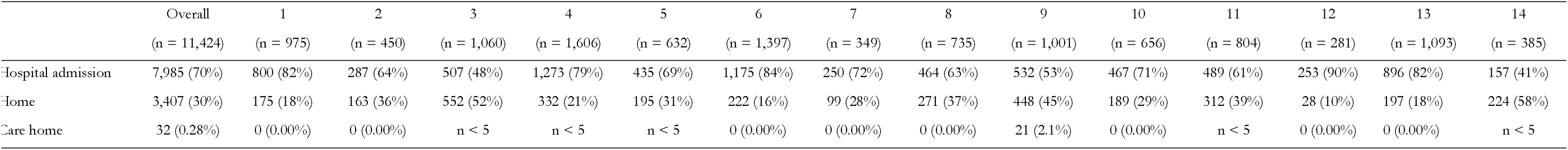
Proportion of patients moving into a temporary stay facility after hospital discharge or directly from their homes or a care home, overall and stratified by municipalities.

**Supplementary Table 3.** The 10 most frequently reported primary diagnoses (codes and names) for hospital admissions of patients discharged to temporary stay facilities.

| Reason for hospital admission, diagnosis code | Reason for hospital admission, diagnosis name | Proportion of hospitalized patients, n (%)<br>(n = 7,985) |
| --- | --- | --- |
| Z508 | Rehabilitation, other | 520 (6.5) |
| S720 | Fracture of head and neck of femur | 316 (4.0) |
| J189 | Pneumonia, unspecified organism | 312 (3.9) |
| S721 | Pertrochanteric fracture | 311 (3.9) |
| Z509 | Rehabilitation, unspecified | 285 (3.6) |
| I639 | Cerebral infarction, unspecified | 223 (2.8) |
| R296 | Repeated falls | 162 (2.0) |
| E869A | Dehydration | 144 (1.8) |
| N390 | Urinary tract infection, site not specified | 113 (1.4) |
| N300 | Acute cystitis | 91 (1.1) |

**Supplementary Table 4.**
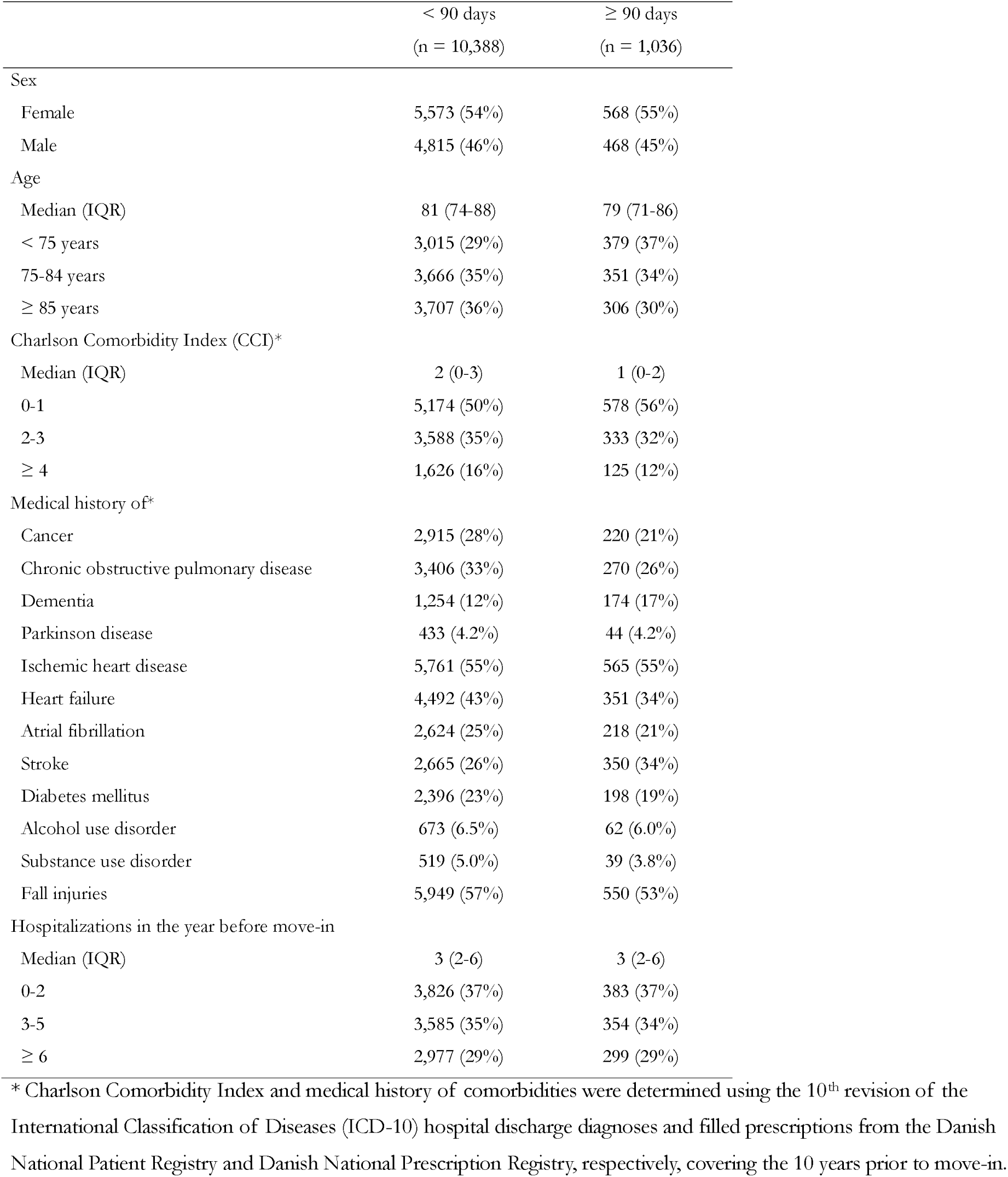
Baseline characteristics of patients moving into temporary stay facilities in 14 Danish municipalities from 2016 to 2023 stratified by temporary stay length.

**Supplementary Figure 1.**
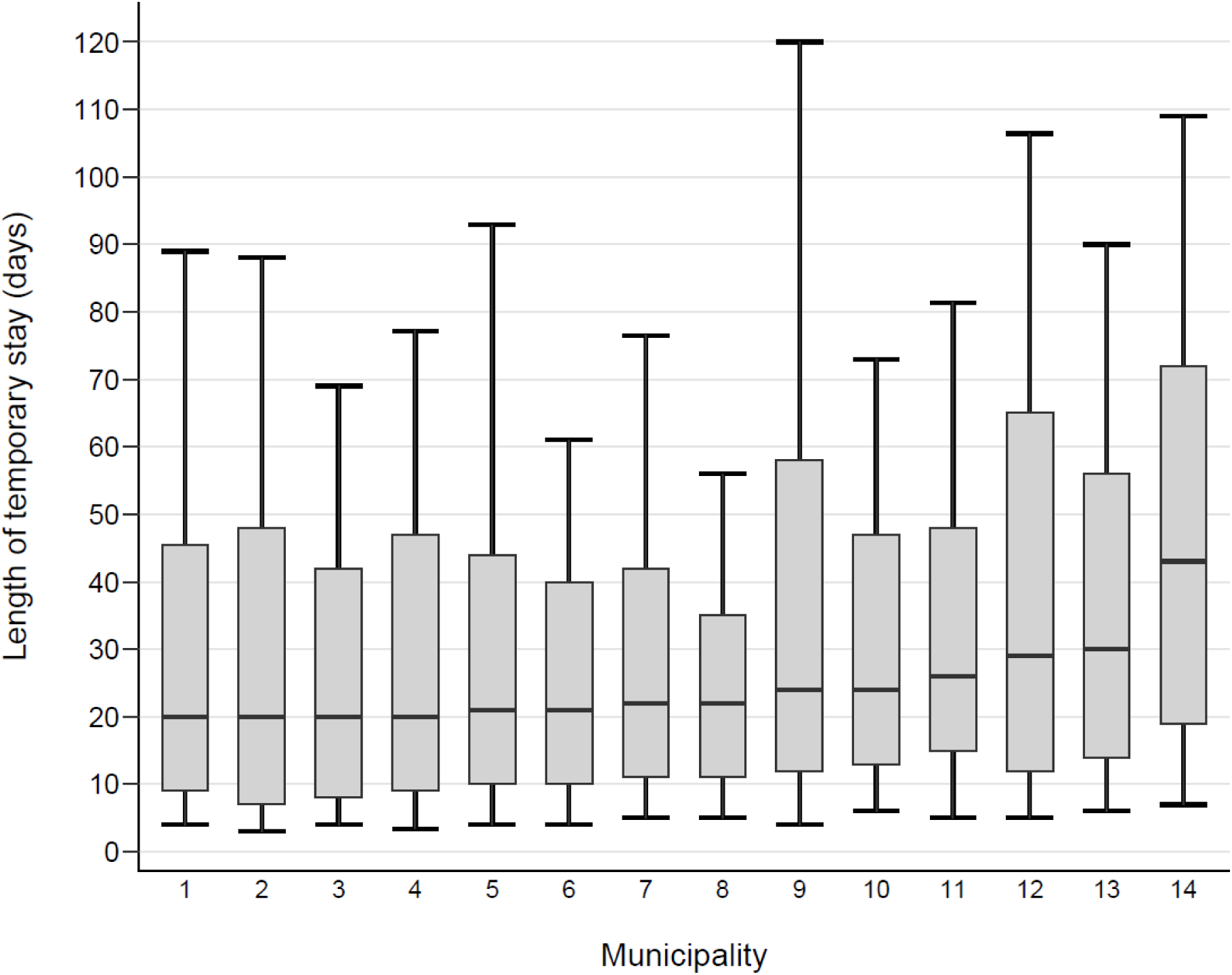
Boxplot of temporary stay lengths by municipality. Each box represents the interquartile range (IQR), with the bottom and top edges indicating the first and third quartiles, respectively, and the midline indicating the median. The whiskers extend to the 10th and 90th percentiles.

**Supplementary Figure 2.**
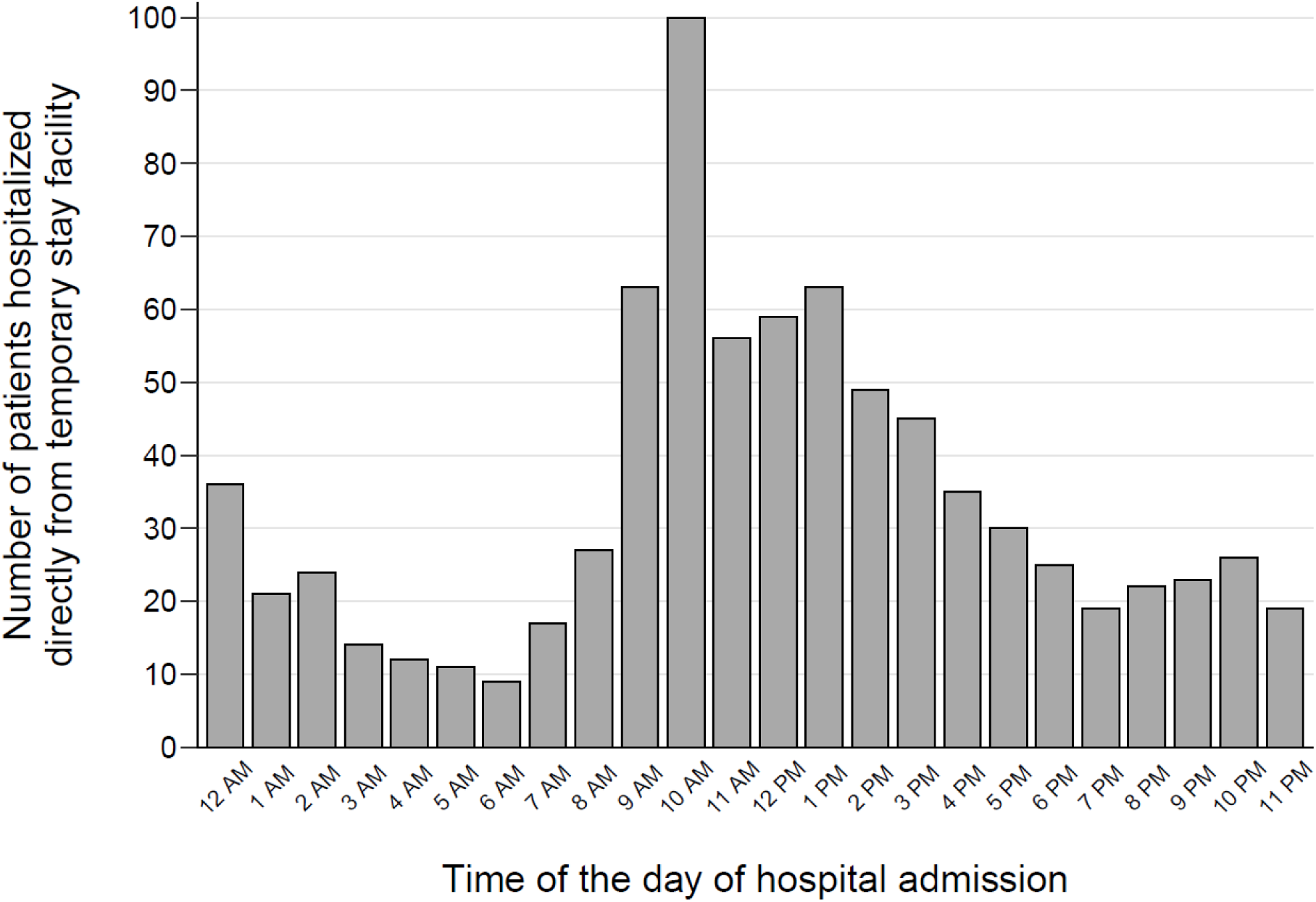
Distribution of hospital admissions by hour for patients admitted directly from temporary stay facilities.

**Supplementary Figure 3.**
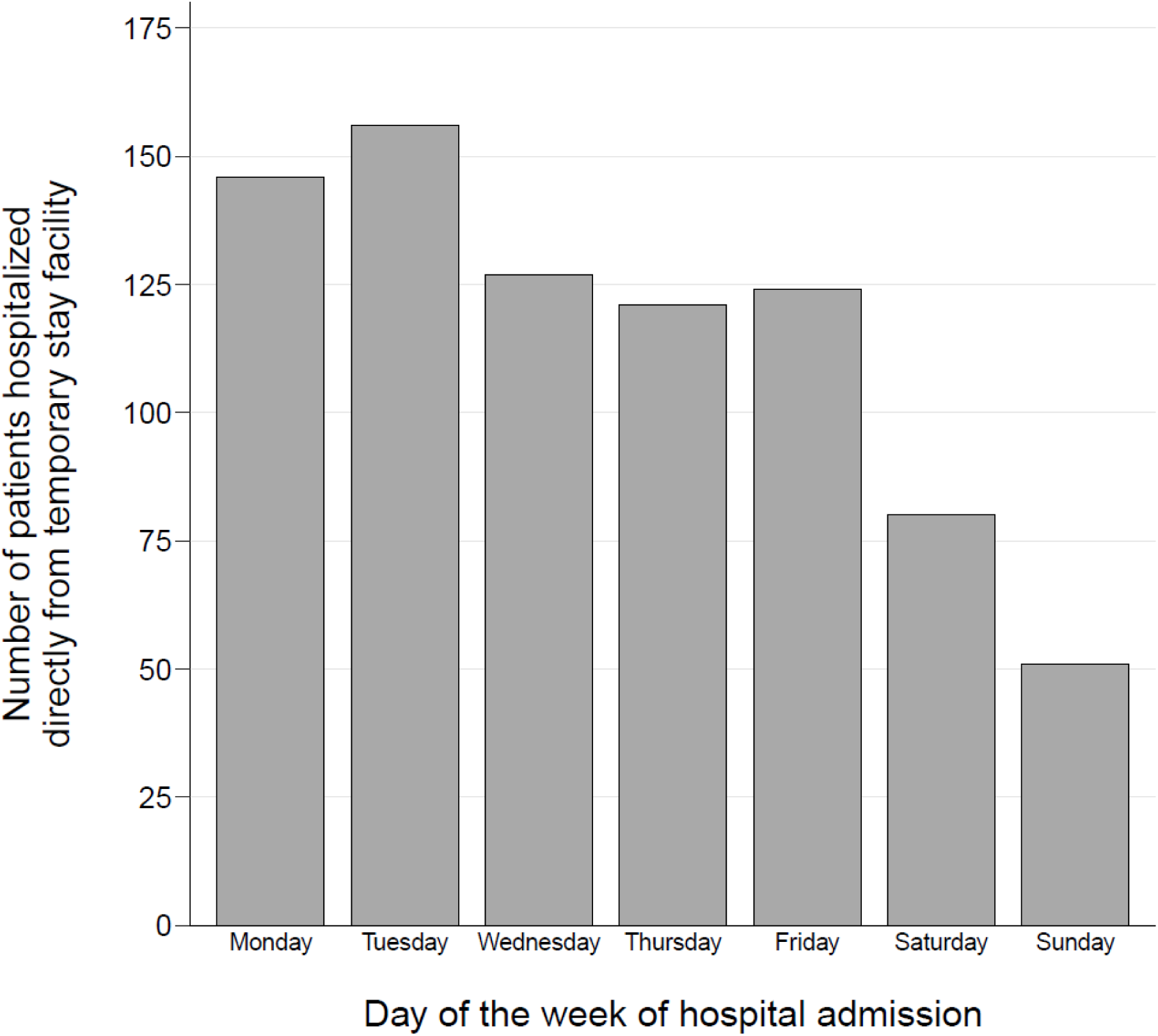
Distribution of hospital admissions by day of the week for patients admitted directly from temporary stay facilities.

**Supplementary Table 5.** The 10 most frequently reported primary diagnoses (codes and names) for hospital admissions of patients hospitalised directly from temporary stay facilities.

| Reason for hospital admission, diagnosis code | Reason for hospital admission, diagnosis name | Proportion of hospitalized patients, n (%)<br>(n = 805) |
| --- | --- | --- |
| J189 | Pneumonia, unspecified organism | 55 (6.8) |
| Z515S | Need for specialised palliative care | 49 (6.1) |
| Z016 | Radiological examination | 34 (4.2) |
| Z039 | Observation for suspected disease or condition, unspecified | 26 (3.2) |
| R060 | Dyspnea | 23 (2.9) |
| Z508 | Rehabilitation, other | 21 (2.6) |
| A419B | Urosepsis | 14 (1.7) |
| E869A | Dehydration | 14 (1.7) |
| A499 | Bacterial infection, unspecified | 14 (1.7) |
| J159 | Bacterial pneumonia, unspecified | 11 (1.4) |

**Supplementary Table 6.** The 10 most frequently reported primary diagnoses (codes and names) for hospital admissions of patients hospitalised directly from temporary stay facilities on weekdays.

| Reason for hospital admission, diagnosis code | Reason for hospital admission, diagnosis name | Proportion of hospitalized patients, n (%)<br>(n = 674) |
| --- | --- | --- |
| Z515S | Need for specialised palliative care | 49 (7.3) |
| J189 | Pneumonia, unspecified organism | 42 (6.2) |
| Z016 | Radiological examination | 34 (5.0) |
| Z039 | Observation for suspected disease or condition, unspecified | 22 (3.3) |
| Z508 | Rehabilitation, other | 21 (3.1) |
| R060 | Dyspnea | 16 (2.4) |
| A419B | Urosepsis | 12 (1.8) |
| A499 | Bacterial infection, unspecified | 12 (1.8) |
| J159 | Bacterial pneumonia, unspecified | 10 (1.5) |
| E869A | Dehydration | 9 (1.3) |

**Supplementary Table 7.** The 10 most frequently reported primary diagnoses (codes and names) for hospital admissions of patients hospitalised directly from temporary stay facilities on weekends.

| Reason for hospital admission, diagnosis code | Reason for hospital admission, diagnosis name | Proportion of hospitalized patients, n (%)<br>(n = 131) |
| --- | --- | --- |
| J189 | Pneumonia, unspecified organism | 13 (9.9) |
| R060 | Dyspnea | 7 (5.3) |
| E869A | Dehydration | 5 (3.8) |
| R509 | Fever, unspecified | n < 5 |
| Z039 | Observation for suspected disease or condition, unspecified | n < 5 |
| A419 | Sepsis, unspecified organism | n < 5 |
| J449 | Chronic obstructive pulmonary disease, unspecified | n < 5 |
| N300 | Acute cystitis | n < 5 |
| Z038 | Observation for other suspected diseases and conditions | n < 5 |
| B342A | COVID-19 infection without localization | n < 5 |

**Supplementary Table 8.**
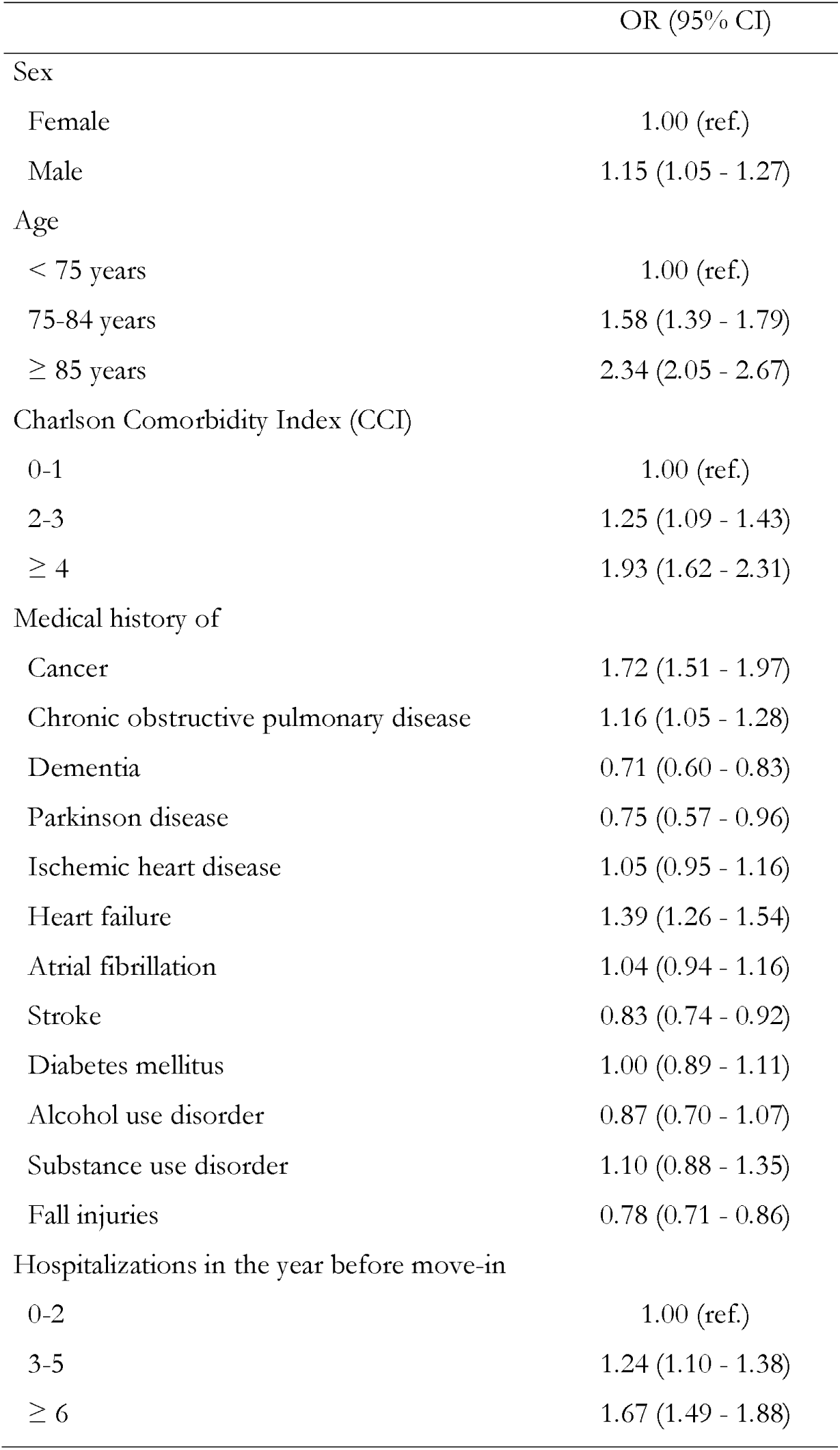
Baseline predictors of 90-day mortality after moving into a temporary stay facility.

## Appendix A – Definition of comorbidities

**A-table 1.** ICD-10 and ATC codes used to define comorbidities in Table 1.

Appendix A – Definition of comorbidities **A-table 1.** ICD-10 and ATC codes used to define comorbidities in Table 1
| Comorbidity | ICD-10 code | ATC code |
| --- | --- | --- |
| Cancer | C D45 D46 D471 D473 D474<br>D475 |  |
| COPD | J40 J41 J42 J43 J44 | R03 |
| Dementia | F00 F01 F02 F03 F1073 F1173<br>F1273 F1373 F1473 F1573<br>F1673 F1873 F1973 | N06D |
| Parkinson disease | F023 G20 G21 G22 | N04BA |
| Ischemic heart disease | I20 I21 I22 I23 I24 I25 | N02BA C01DA B01AC |
| Heart failure | I099A I110 I130 I132 I50 | C03C |
| Atrial fibrillation | I48 |  |
| Stroke | I60 I61 I62 I63 I64 I69 |  |
| Diabetes mellitus | E10 E11 E13 E14 | A10 |
| Alcohol use disorder | F101 | N07BB |
| Substance use disorder | F11 F12 F13 F14 F15 F16 F17<br>F18 F19 | N07BC |
| Fall injuries | S00 S01 S02 S06 S10 S12 S20<br>S21 S22 S30 S31 S32 S40 S41<br>S42 S50 S51 S52 S70 S71 S72<br>S80 S81 S82 S430 S431 S432<br>S530 S531 |  |

*ICD-10 = 10<sup>th</sup> revision of the International Classification of Disease; ATC = Anatomical Therapeutic Chemical; COPD = Chronic Obstructive Pulmonary Disease*

